# COVID-19 Vaccine Hesitancy in India: An Exploratory Analysis

**DOI:** 10.1101/2021.09.15.21263646

**Authors:** Sandip K. Agarwal, Maharnab Naha

## Abstract

Vaccine hesitancy is context specific and varies over time and space. Therefore, strategies to tackle vaccine hesitancy based on evidence from high income countries are unlikely to serve the purpose adequately in LMICs. We use district level evidence on COVID-19 vaccine uptake rates from an LMIC - India to provide evidence of COVID-19 vaccine hesitancy. We argue that vaccination rates during the different phases of COVID-19 vaccination across the districts is likely to be related to vaccine hesitancy. Districts with larger rural population and lower literacy rates had lower vaccination rates. High past child immunization rates were positively correlated with COVID-19 vaccination uptake. Across the four phases of vaccination drive, vaccine hesitancy was the highest during the third phase of the vaccination drive, and therefore the above correlations were strongest during the third and the fourth phase. Measures of family planning indicators too seem to be correlated with vaccine uptakes during the third phase which indicate the regions most susceptible to vaccine hesitancy.

## Introduction

Herd immunity is the key to minimize disruptions caused due to the pandemic. Sooner the population acquires herd immunity, sooner human life and activity can be restored to normal. However, the natural process of building up herd immunity can be slow, which would mean that the pandemic is prolonged. Therefore, acquiring herd immunity through mass inoculation against COVID-19 had been suggested as a priority target for policy makers throughout the world to acquire herd immunity in a sufficient proportion of population (Randolph and Barreiro, 2020).

Scientists have developed vaccines for COVID-19 at unprecedented speed. However, availability of a vaccine does not imply that people will be willing to get inoculated. The WHO Strategic Advisory Group of Experts (SAGE) Working Group on Vaccine Hesitancy concluded that *vaccine hesitancy refers to delay in acceptance or refusal of vaccination despite availability of vaccination services* (MacDonald et al., 2015). In 2019, the World Health Organization (WHO) declared vaccine hesitancy as one of the top ten threats to public health.

Vaccine hesitancy can be one of the greatest hurdles in achieving herd immunity. A successful and an effective vaccination campaign requires much more than the availability of a safe and effective vaccine. This includes identifying sections of the population with lower vaccine confidence or uptake rates and identifying the drivers of vaccine hesitancy. If vaccine hesitancy is timely addressed, this can avert an adverse public health outcome (Larson, 2015b). Empirically grounded models of vaccine hesitancy and vaccine acceptance viz. 5Cs and 5As exist based on evidence from high income countries (Machingaidze, S., & Wiysonge, 2021). However, similar research efforts towards identifying and addressing vaccine hesitancy are lacking for the low income and middle income countries (LMICs). Evidence based research surrounding vaccine hesitancy in the context of LMICs requires attention to increase vaccination uptake by fighting vaccine hesitancy in the current pandemic (Simas & Larson, 2021).

COVID-19 vaccination is the lowest among the LMICs as most LMICs are relying on other vaccine producing nations for the supply of COVID-19 vaccines. However, even if vaccines are available in sufficient supply, vaccine hesitancy can potentially lower vaccine uptake. Among the LMICs, India is one of the few countries that developed and produced vaccines at home. Although the COVID-19 vaccine roll out has been slower than expected (Pandey et al. 2021), compared to the other LMICs a significant proportion of the Indian population have been vaccinated. Evidence on vaccine uptake and vaccine hesitancy from India’s COVID-19 vaccination drive can serve as an important lesson for vaccination campaigns in India and other LMICs and better public health outcomes in the future.

In this article we provide district level trends and heterogeneity in the COVID-19 vaccination rates in India. Based on district level vaccination rates in India, we find that there are significant differences in vaccine uptake between the rural and the urban areas with relatively lower vaccination uptake in the rural areas. We argue that vaccine hesitancy contributed to lower COVID-19 vaccinationation rates in India and identify the potential socio-economic factors and past health and family planning indicators that are correlated with COVID-19 vaccination rates. Our paper is organized as follows. Section 2 lays down a historical narrative of prominent anti-vaccine sentiments in India. Section 3 builds the timeline of Covid-19 vaccination drive by India. Section 4 describes the data and provides evidence of spatial and temporal heterogeneity in the COVID-19 vaccination rates. Section 5 discusses the evidence and the possible causes of vaccine hesitancy. Section 6 concludes.

## Background

In the 1970s India set up the immunization programme that was later renamed as the Universal Immunization Program (UIP). Anti-vaccine sentiments in India had never been an organized anti-vaccine movement unlike the west. Prior to the UIP during British India and after independence few prominent Indian leaders who had mass appeal among the people were apprehensive of safety and efficacy of the vaccines or questioned the compatibility of vaccines with their religious beliefs. The UIP met with criticism from a section of elitists on account of being a relatively costly program for a poor country like India (Brimnes, 2017). However, since its inception UIP has led to successfully eliminating polio and maternal and neonatal tetanus from India by 2015 (MOHFW, GOI). Besides these, there have been few lone episodes of vaccine hesitancy in relatively smaller pockets in India that caught international attention. In the early 2000s resistance against the polio vaccine surfaced in some districts of Uttar Pradesh and Bihar following rumors of sterilization among the marginalized communities (Larson, 2015a). Another instance of anti-vaccine sentiment was the introduction of Hib vaccine in the UIP in 2010 due to which a larger rollout of the Hib vaccine was suspended (Mudur, 2010; Saxena et al., 2010).

Vaccine hesitancy is context specific and can evolve with changing norms in the society and rapid progress in information and communication. Adult immunization is not popular in the LMICs including India unlike the west where there is some familiarity with adult vaccination. However, the success and the size of the UIP in India built unrealistic confidence in the potential uptake of vaccines (Biswas, 2020; Pinnamaneni and Seshasayee, 2021). While all attention was focussed on supply of vaccines, little attention was given to mobilize the demand for vaccines (Chowdhury et al., 2021). The availability of a vaccine does not mean that people will rush to get inoculated. Introduction of a new vaccine demands rigorous research surrounding psychological, social and political aspects to assess public trust in the vaccine as much as it demands scientific rigorous evidence on safety and efficacy of the vaccine (Black and Rapoulli, 2010; Larson et al., 2011). Therefore, a multi-disciplinary approach is needed by bringing researchers together from behavioral sciences, economics, sociology and other social sciences that could address questions of reduction in vaccine hesitancy and improvement of immunization outcomes (Black and Rapoulli, 2010).

### Covid-19 Vaccination in India

Covid-19 vaccination drive for adults in India began on January 16, 2021 with two double-dose approved vaccines. The vaccination drive was rolled out in four phases which is depicted in figure 1. Phase-1 between January 16 to February 28 included vaccination of all frontline workers, who were actively involved in containing the spread of the pandemic. Frontline workers included health workers, security staff etc. Phase-2 of the vaccination campaign scheduled during March 2021 was opened to all aged 60 years and above and individuals aged 45 years and above with comorbidities like hypertension, diabetes, HIV infection etc. During phase-3 in the month of April all individuals aged 45 years and above were eligible for vaccination. The fourth phase of vaccination that began on May 1 was opened to all adults 18 years and above.

**Figure 1:**
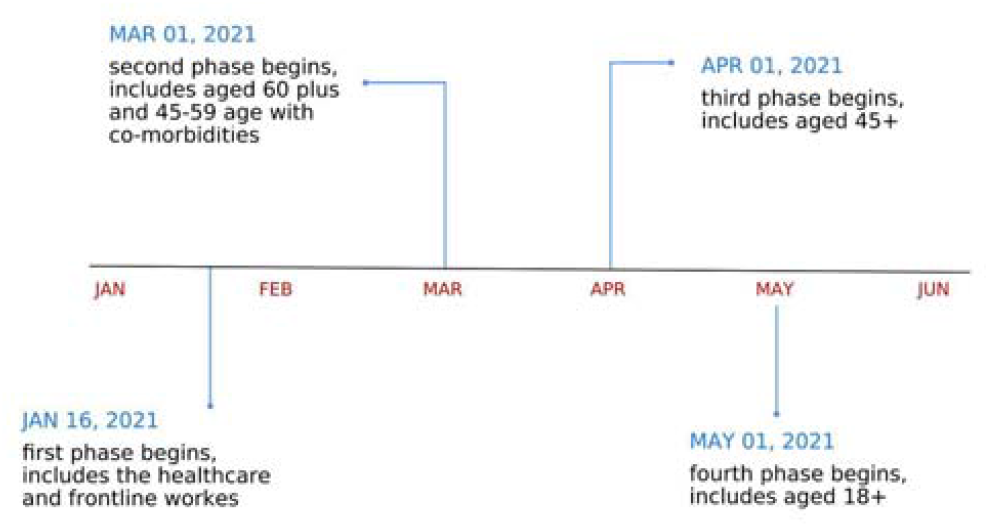
Timeline for Covid-19 vaccination drive in India

Vaccination against COVID-19 was voluntary for all. This meant that individuals could delay in accepting the vaccine or refuse to get vaccinated in spite of the availability of a vaccine. Vaccine confidence surveys can identify vaccine hesitancy spots. In the past Vaccine Confidence Project (VCP) and several other studies have surveyed attitudes towards vaccination globally (Larson, 2015a; Larson, 2015b; Figueredo et al., 2016; Figueredo et al., 2020; Sturgis et al., 2021). Similar surveys were conducted in several countries to assess the confidence in COVID-19 vaccine. If reasons for vaccine hesitancy can be timely addressed, this could increase uptake of vaccination, when a vaccine becomes available (Dubé & MacDonald, 2020; Sallam et al., 2021).

In figure 2, we have compiled and plotted the attitude towards COVID-19 vaccine in India as measured in different surveys between July 2020 and July 2021. More details about the surveys used can be accessed in the appendix. While there is considerable heterogeneity in vaccine hesitancy as obtained from various surveys, existence of vaccine hesitancy cannot be denied. Vaccine hesitancy for COVID-19 vaccines in India appears to be clustered around 60%. Surveys that have indicated lower vaccine hesitancy are smaller in sample size and less representative due to higher representation of respondents from urban areas in India (Lazarus et al., 2021). Sub-national evidence of vaccine hesitancy for COVID-19 vaccination in India has been reported from different regions and specific groups in the population (Jain et al., 2021; Kumari et al., 2021; Noronha et al., 2021; Roy chowdhury et al., 2021; Wagner, 2021)

**Figure 2:**
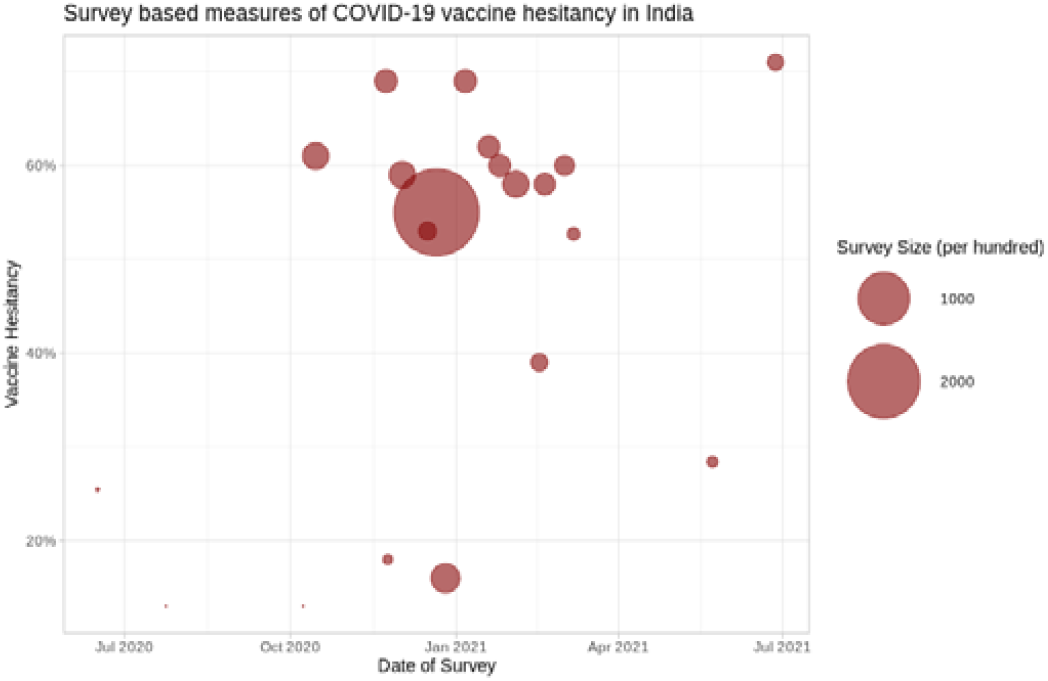
Vaccine Hesitancy as measured in different vaccine surveys (Authors compilation)

Figure 3 and 4 plots the daily administered COVID-19 vaccine doses and daily COVID-19 infections and related deaths respectively. Phased roll out of COVID-19 vaccines in India prioritized the most vulnerable to get vaccinated first based on age and medical history. As per the demographic structure of the Indian population, proportions of the population eligible for vaccination during phase 3 and 4 were significantly higher than those eligible under phase 1 and 2 (SRS, 2016). However, as we can observe in figure 3 there was a consistent decline in daily COVID-19 vaccine doses administered during phase 3 and initial month of the phase 4 of the vaccination drive. This decline in administered vaccine doses coincided with the peak of the second wave of covid infection as it can be seen in figure 4. September 2020 and April 2021 were the peak months of the first and the second wave of the pandemic in India respectively with the latter being more severe (Lancet, 2021).

**Figure 3:**
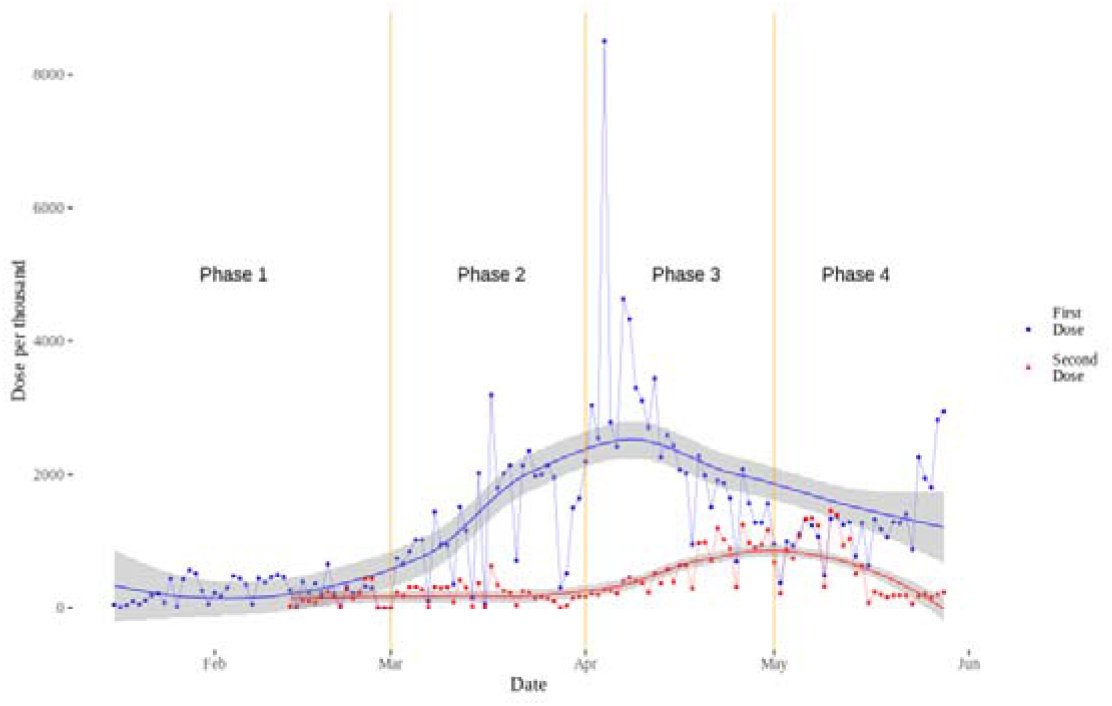
Daily COVID-19 vaccination in India

**Figure 4:**
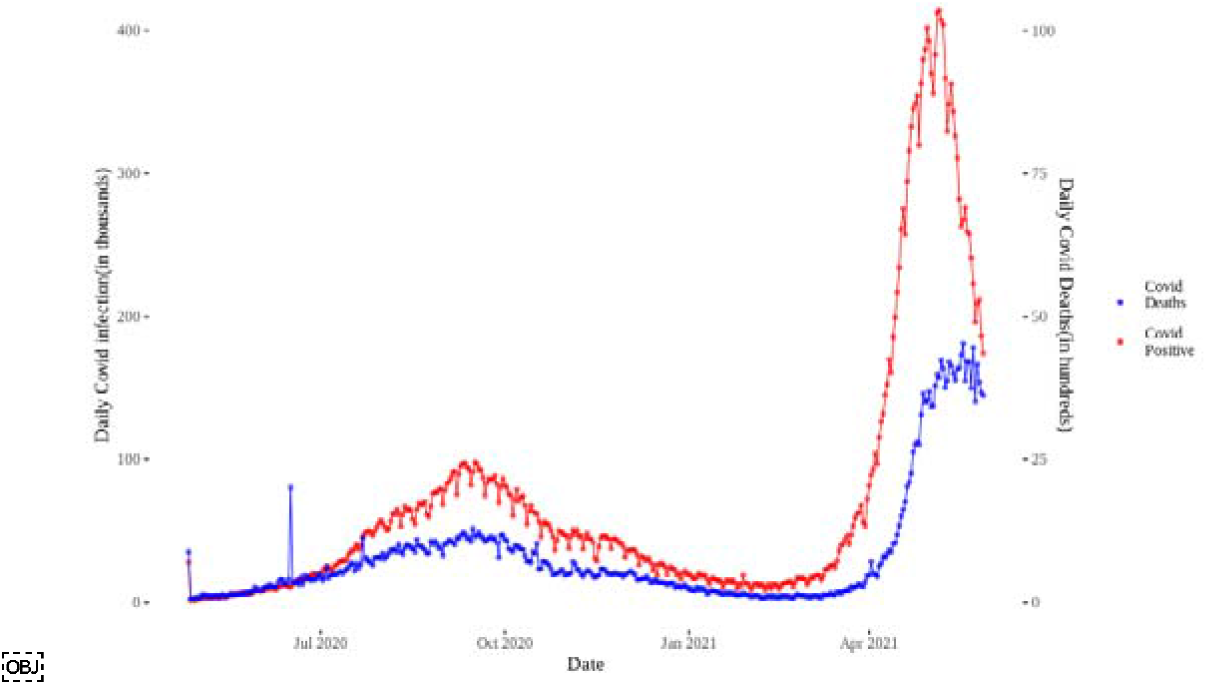
Daily COVID-19 infections and COVID-19 related deaths in India

The decline in administered vaccine doses during the third and the fourth phase of vaccination drive could be due to short supply of vaccines or low demand for vaccines or both. While limited vaccine supply has affected the daily administered vaccine doses, there is no evidence towards consistent decline in total supply of vaccines during the entire phase 3 and initial month of phase 4. Although data on the production and stock of COVID-19 vaccines in India is not available to researchers yet, we are not aware of any evidence that can support the trend of observed decline in daily vaccine doses during the third and fourth phase of the vaccination drive. However, it is possible that individuals faced physical and psychological barriers in accessing the vaccines in spite of their availability. Localized lockdowns might have reduced mobility or the second wave of COVID-19 infection might have created fear of contracting COVID-19 that inhibited people to visit vaccination centres. Nonetheless, these are considered as antecedents to vaccine hesitancy or vaccine acceptance as explained by 5A and 5C models of vaccination (Thompson et al., 2016; Betsch et al., 2018). Therefore, we believe that vaccine hesitancy significantly contributed to the observed decline in the daily administered doses of COVID-19 vaccines during third and the fourth phase of the COVID-19 vaccination campaign in India.

COVID-19 vaccines in India and elsewhere are mostly two shot vaccines. High efficiency requires both doses of COVID-19 vaccines to be administered that can provide protection against fatal infection. As the second wave of COVID-19 infection picked up, India saw a high rate of COVID infection and deaths among the elderly people and people with comorbidities. Elderly and the people with comorbidities were the ones who were at high risk of COVID-19 infection, and hence they were prioritized to receive COVID-19 vaccine shots relative to others in the phase wise vaccine rollout. Many among them who were vaccinated with the first dose contracted COVID-19 infection and some of them died as the first dose alone was not meant to build sufficient immunity to fight severe COVID infection. However, the myth developed particularly in the rural areas that covid vaccines were causing deaths and illness. This too likely contributed to vaccine hesitancy during the third and the fourth phase of the vaccination drive. Researchers have termed this phenomenon as “Co-incidence dragon” [Post hoc ergo propter hoc: after this, therefore because of this] which is the cognitive flaw in reasoning that stems from natural human desire to find order and predictability in random data (Jacobson et al., 2007; MacDonald et al., 2012).

## Data and Methodology

We have argued in the previous section that the decline in the daily vaccination during the third and the fourth phase is not solely due to short supply of vaccine but also due to vaccine hesitancy. In this section we use exploratory analysis to analyze the heterogeneity in COVID-19 vaccination uptake across districts in India through the four phases of the vaccination drive. A district is an administrative unit lower than the states analogous to a county in the US. As of 2021 there are a total of 718 districts in India.

Daily vaccination data for districts is obtained from the Cowin portal of Government of India (https://www.cowin.gov.in/), which is further aggregated to get the total number of first doses of vaccines administered during each phase. Fourth phase of vaccination in our data only includes the first dose of vaccines administered between May 1st and May 31st. We calculate district level vaccination rate for the first dose of vaccination by finding the population vaccinated in each district as a percentage of district level population. While the primary source of district level population data is from the Indian Census 2011, we use district level population data from Wang et al. (2020). Wang et al. (2020) have estimated population data for 2020 for the districts in India based on the census data. The advantage of using estimated district level population data for the year 2020 is that the risk of overestimating vaccination rates for the district is minimized which is likely to be the case with the use of 2011 census data.

Figure 5 and 6 plots the district level vaccination rate for the first dose against the percentage of rural population and literacy rate in a district during the four phases of COVID-19 vaccination campaign. The fourth phase data includes only the first month of the fourth phase vaccination. Data on percentage of rural and literate population in a district was obtained from the 2011 Census of India. For figures 5 and 6 the districts from 2021 COWIN portal were mapped to the districts of 2011 Census. The above plots also represent the linear fit for each phase.

**Figure 5:**
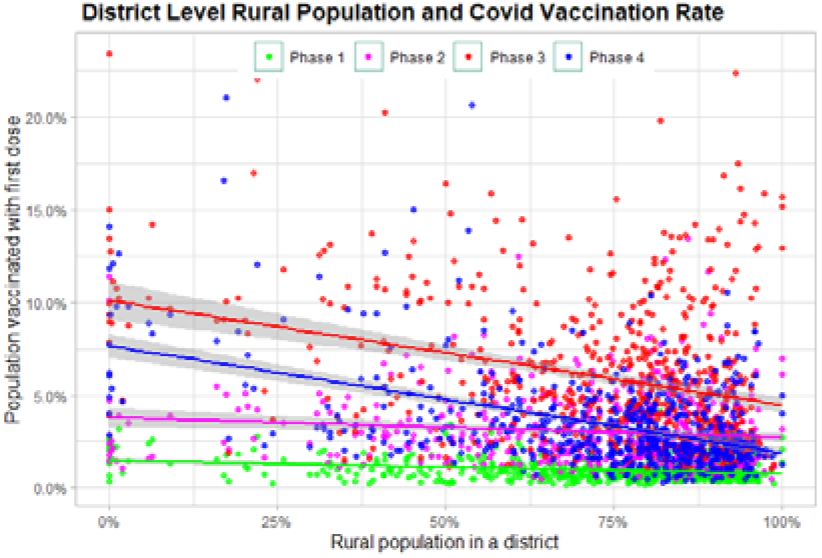
Rurality and vaccination rate

**Figure 6:**
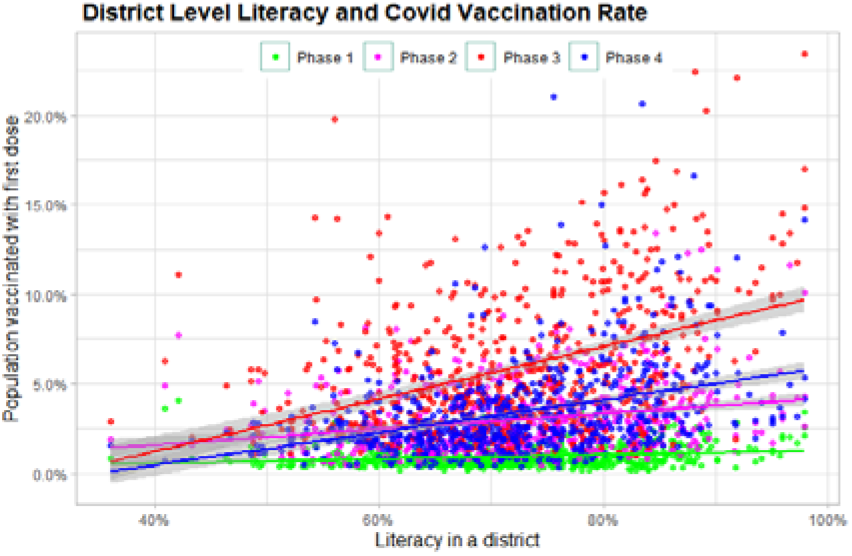
Literacy and Vaccination rates

Similarly, figures 7 and 8 plots the variation in vaccination rates during the four phases across the percentage of Scheduled Castes (SC) and Scheduled Tribes (ST) in each district. SC and ST are the marginal population, who have a history of social exclusion faced by them. Figure 9 plots the vaccination rates across the percentage fo muslim poulation in a district. Muslims are a religious minority in India. The state of Jammu & Kashmir (J&K) in India was an outlier in vaccination rate as it had high vaccination rates (The Print, 2021; Hindustan Times, 2021). Several districts in J&K had more than 90% population as muslims. Therefore, we recreated another scatter plot as in the right panel of figure 9 by excluding the districts of the state of J&K.

**Figure 7:**
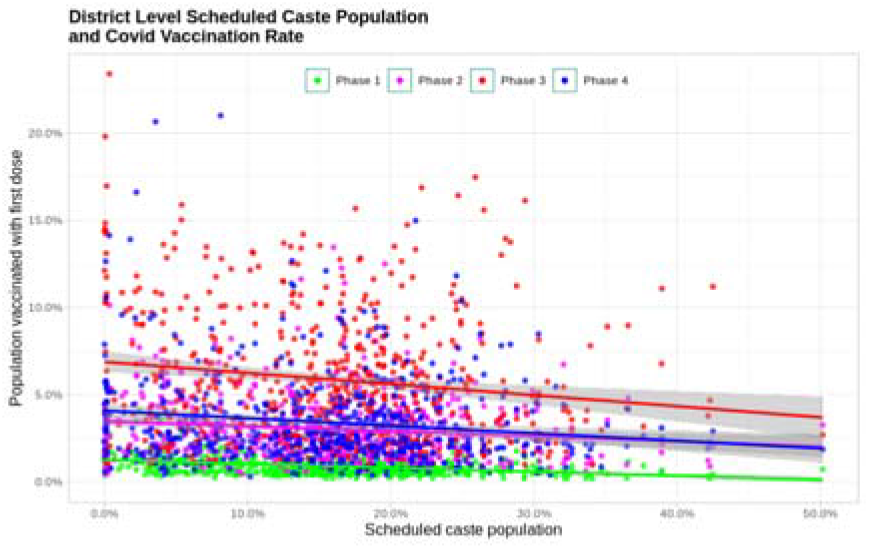
SC population and vaccination rate

**Figure 8:**
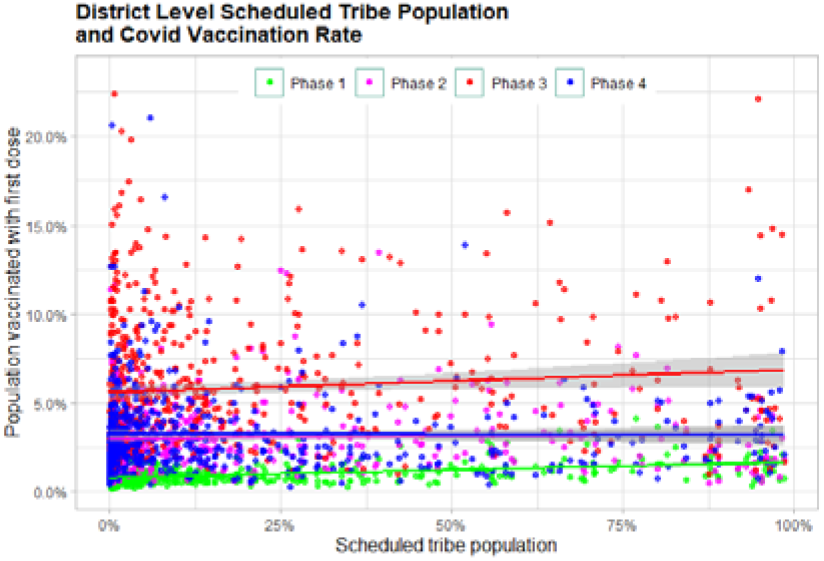
ST population and vaccination rate

**Figure 9:**
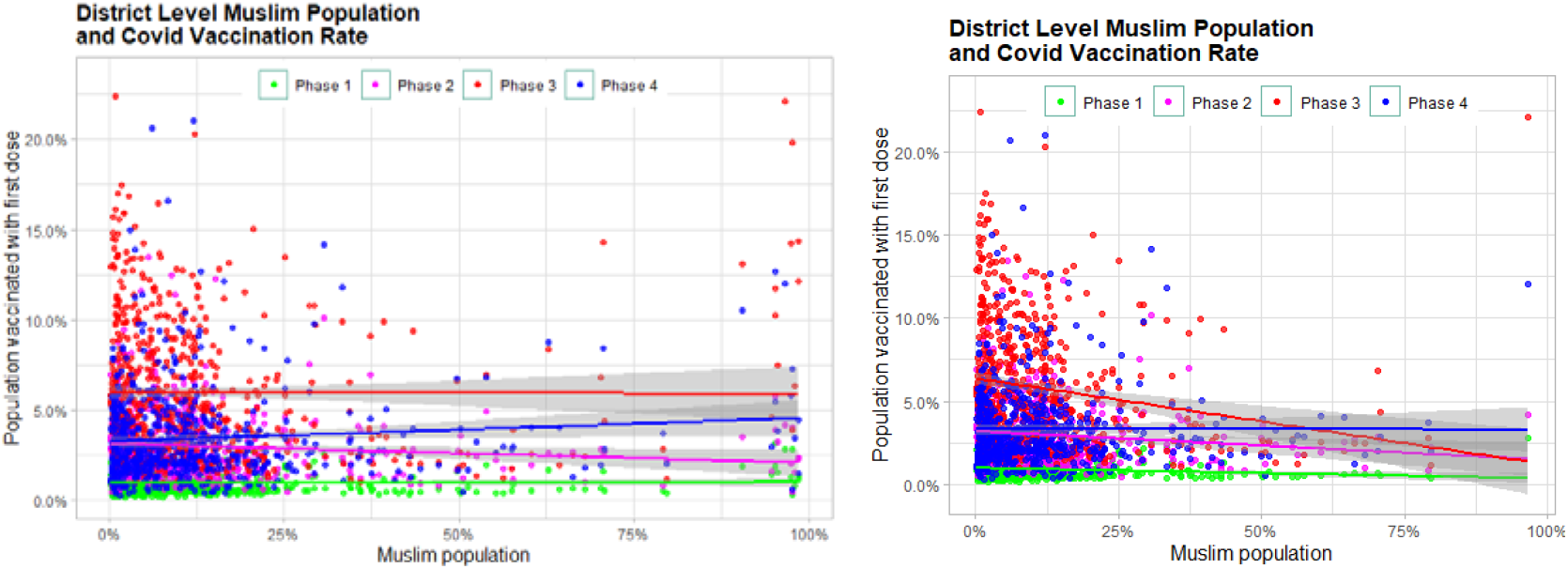
Muslim population and vaccination rate (including and excluding Jammu and Kashmir)

We use information on district level survey data on past child vaccination rates and adoption of family planning methods from the National Family and Health Survey (NFHS) data to find the potential correlation with district level COVID-19 vaccination rates across the four phases. We use the fourth and the fifth rounds of the NFHS data pertaining to the years 2015-16 and 2019-20 respectively. NFHS-5 data was released at the end of 2020 which did not include all the states as the survey data collection was affected by the pandemic. Hence, our plots for the NFHS-5 include only the states for which the data was available. Districts from the COWIN portal were mapped to the districts in each round of NFHS data for plotting the graphs.

A child is considered fully immunized if the child has been inoculated with three doses of diphtheria-tetanus-pertussis (DTP) vaccine, three doses of polio vaccine, one dose of Bacillus Calmette–Guérin (BCG) vaccine, and one dose of measles vaccine. Heterogeneity in the child immunization rates could potentially reflect general attitude towards vaccines, hence could predict regions of vaccine hesitancy. Figure 10 plots the district level COVID-19 vaccination rates across district level full immunization rates of children from both the fourth and the fifth rounds of the NFHS data in the left and the right panel respectively. The left panel corresponding to 2019-20 years has a lesser number of districts as compared to the right panel as the NFHS-5 data was released partially that did not include districts from all the states.

**Figure 10:**
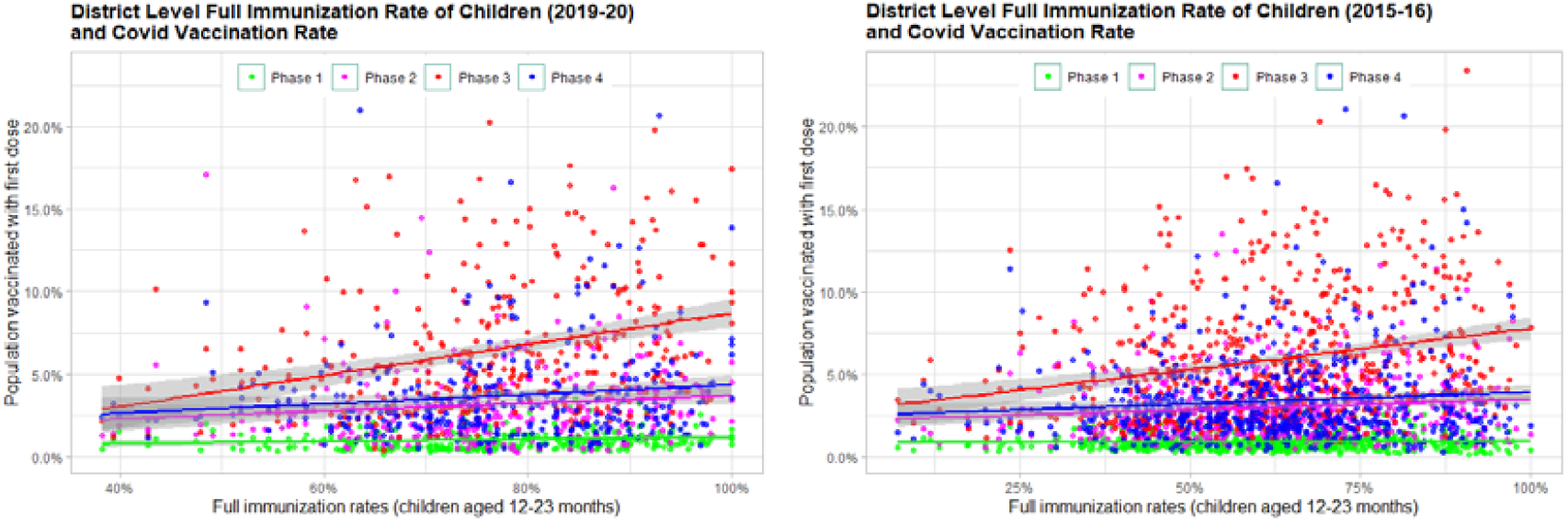
District level Covid-19 vaccination rates and children immunization rates

Similarly, acceptance and adoption of family planning methods could be another predictor of vaccine hesitancy. Evidence from several parts of the world suggest that vaccine hesitancy is associated with myths surrounding fertility concerns. Therefore, we also find the correlation of district level COVID-19 vaccination rates with adoption of family planning practices. Figure 11 plots the district level vaccination rates across the percentage of people in a district adopting any method of family planning for the rounds four and five of NFHS data. Among the family planning methods, the most popular method was female sterilization. Therefore, we also plot graphs that correlate COVID-19 vaccination rates with female sterilization rates as in figure 12. The left and the right panels in figure 12 correspond to 2019-20 and 2015-16 rounds of NFHS data respectively. Figure 13 left and right panel plots the fourth and fifth round of NFHS data respectively for total unmet need for family planning on the horizontal axis with the COVID-19 vaccination rates on the vertical axis. Unmet need for family planning in a district measures the percentage of married women between the age of 15 to 49 years, who wish to adopt a family planning method but are currently not using any.

**Figure 11:**
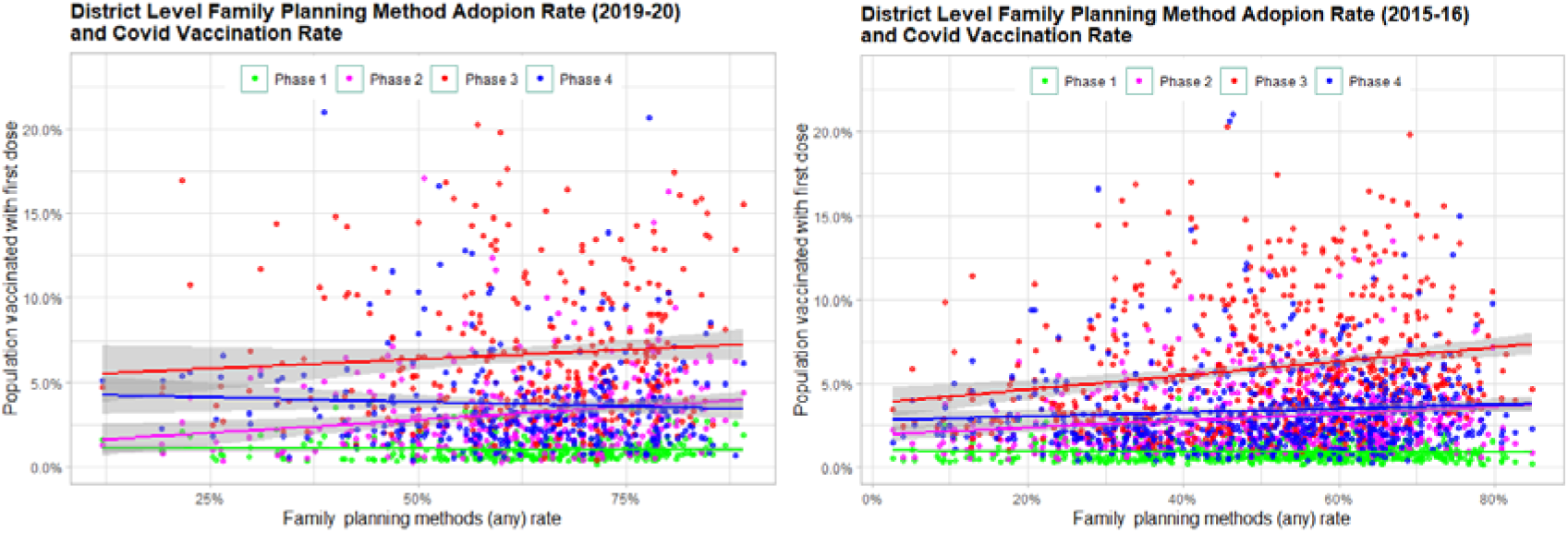
District level Covid-19 vaccination rates and Family Planning Adoption rates

**Figure 12:**
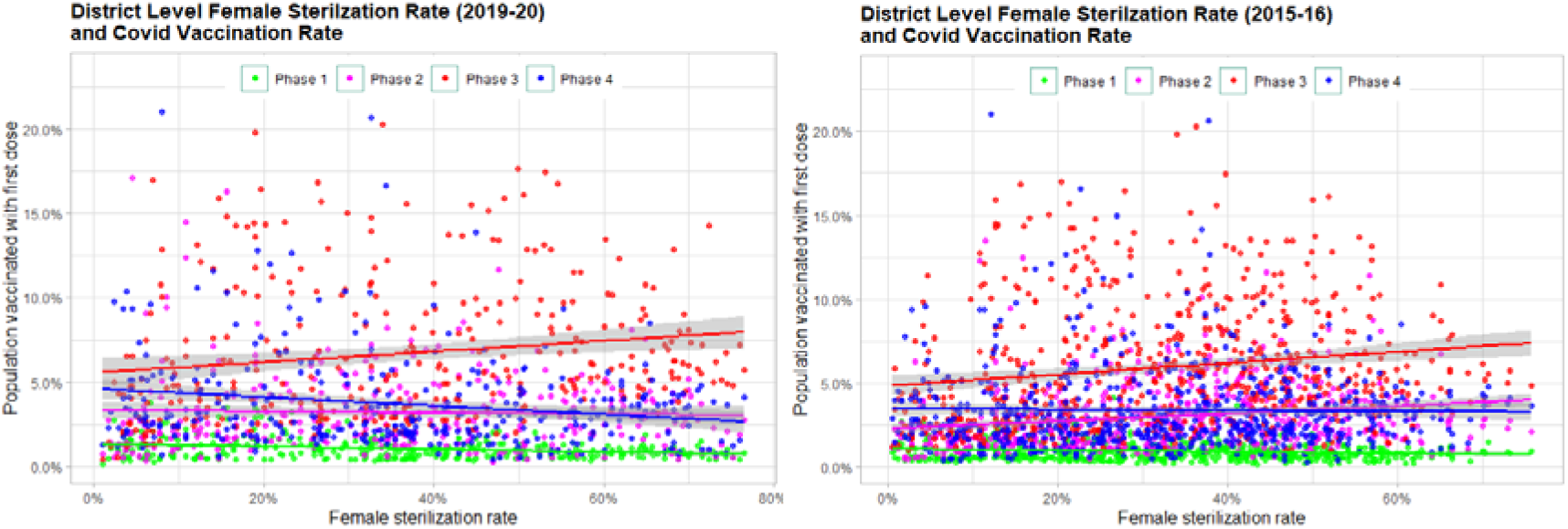
District level Covid-19 vaccination rates and Female sterilization rates

**Figure 13:**
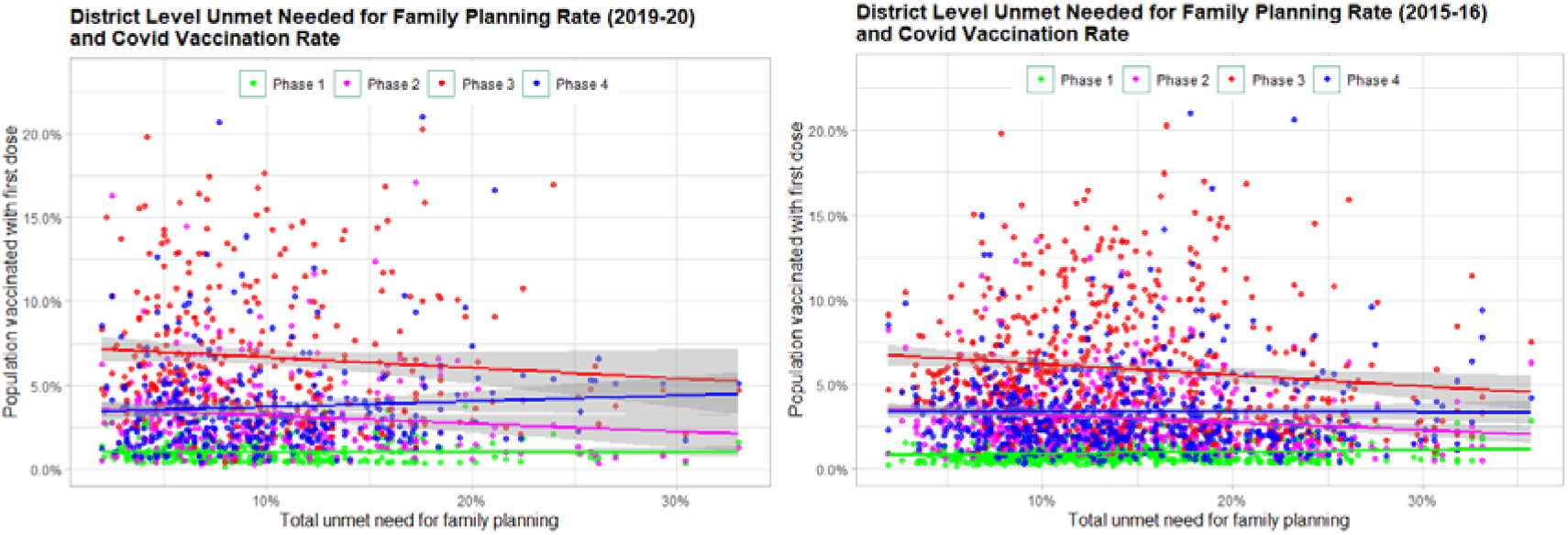
District level Covid-19 vaccination rates and Unmet need for family planning

## Discussion

A successful vaccination campaign can potentially mitigate the costs of the pandemic by inducing herd immunity required for social and economic activities at the pre-pandemic level. However, a successful vaccination drive requires much more than the availability of a safe and efficient vaccine. This includes identifying the spots of vaccine hesitancy and timely addressing the factors that give rise to vaccine hesitancy. While research surrounding vaccine hesitancy has been a subject of serious research in high income countries, similar initiatives have been lacking for LMICs (Simas & Larson, 2021; Machingaidze, S., & Wiysonge, 2021). Empirically grounded models of vaccine hesitancy or acceptance like the 5Cs and 5As have been developed based on research and evidence from high income countries (Machingaidze, S., & Wiysonge, 2021). However, vaccine hesitancy is context specific (MacDonald et al., 2015) which vary across population, culture, region. Therefore, vaccination models tailored to LMICs need to be developed to understand vaccination and tackle vaccine hesitancy (Simas & Larson, 2021).

We use district level evidence from India, and find the correlation of district level vaccination rates with socio-economic factors and relevant health and family planning indicators. A successful vaccination campaign requires identifying the spots of vaccine hesitancy or the spots which are mostly likely to develop vaccine hesitancy and address the concerns before the hesitancy spreads to other regions. A comparison of district level vaccination rates across the four phases help us uncover the socio-economic factors, which could be likely predictors of vaccine hesitancy in India. The dip in the daily vaccination numbers observed during the third phase and the early fourth phase of COVID-19 vaccination drive in India (as seen in figure 3) indicates the presence of high vaccine hesitancy as discussed earlier. During this phase, heterogeneity in the district level vaccination rates increased. Therefore, comparison of vaccination rates across different phases of vaccination drive could identify the spots that are most vulnerable to vaccine hesitancy.

Phase 1 of the vaccination drive inoculated all frontline workers which included workers in the health sector and emergency services that had the highest risk of covid infection due to the nature of their job. There is no heterogeneity in the vaccination rates in the first phase across different socio-economic variables in the district. While there has been evidence of vaccine hesitancy among frontline workers in India (Jain et al., 2021; Mehta et al., 2021), getting vaccinated was the best available alternative for most frontline workers due to the high risk of covid infection from their occupation. Moreover, frontline workers were a tiny percentage of the total district population. Therefore, it is unlikely that vaccine hesitancy among frontline workers would correlate with district level demographic and socio-economic factors or any other health indicators.

There has been high heterogeneity in the vaccination rates between the rural and the urban districts. While the evidence points toward low vaccination rates among the rural population relative to the urban population, this gap is evident only during the third and the fourth phase of the vaccination drive, when the vaccine hesitancy was the highest as argued earlier. During the early phases of the pandemic, primarily the urban areas were infected with COVID-19 and therefore had higher awareness about the disease and the vaccine relative to the rural population. However, the rural population was much affected during the second wave of the COVID infection that coincided with the third phase and the early fourth phase of COVID-19 vaccination drive. In the phase wise rollout of the COVID-19 vaccines, the first and the second phase vaccinated the population most vulnerable to COVID-19 infection. The deaths and infections among the people vaccinated with the first dose of COVID-19 vaccine during the second wave of COVID-19 infection led to the belief that vaccines were deadly, which harmed the vaccination drive. However, we cannot deny that this could have been driven due to heterogeneities in the supply of COVID-19 vaccines. If it would have been the case that during the peak of the second wave urban areas were prioritized for vaccine supply over rural areas, it could have driven the disparities between the urban and the rural vaccination rates.

Literacy rate is positively correlated with vaccination rates. Evidence exists from past research that higher levels of schooling were associated with higher vaccine acceptance in general and with respect to Covid-19 vaccines (De Figueiredo et al., 2020; Priya et al., 2020; Lazarus, 2021). Therefore, it is not surprising to find districts that had higher literacy rates also to have higher vaccination rates. However, in the context of Covid-19 vaccination drive in India, there was a linguistic barrier that could have likely strengthened the relationship between literacy rates and covid-19 vaccination uptake. The registration process for booking a vaccination slot was through an online app in English language. According to the 2011 census, only over 10% of the Indian population use English as their first, second or third language and that too it is more prevalent among the urban population. Therefore, linguistic barriers could have significantly contributed to vaccine hesitancy. Behavioral models of vaccination identify linguistic barriers as one of the reasons for vaccine hesitancy, which are classified as Constraint or lack of Convenience under 5C model and lack of Access under 5A model (Betsch et al., 2018; Thompson et al, 2016). The positive relationship between literacy rates and vaccine uptake was more pronounced during the third and the fourth phase of vaccination that coincided with the peak of the second wave of covid infection. This is also indicative of the fact that vaccine hesitancy regions with low literacy rates are most susceptible to vaccine hesitancy.

Past research has indicated lower vaccination rates among the marginalized sections of the society have been reported in the past (Shrivastwa et al, 2015; Shrivastwa et al, 2019; Razai et al., 2021). The marginalized groups in India that consist of SCs and STs have a long history of exclusion. Therefore, inequitable access to the health system and mistrust in the system are possible reasons for lower vaccination rate among the marginalized sections of the society (Shrivastwa et al, 2019; Hunter et al., 2021; Morales & Ali, 2021). Districts with relatively higher concentration of SC population have lower vaccination rates during the second, third and initial month of the fourth phase of the covid-19 vaccination drive. However, we do not see a trend in vaccination rate across the ST population. A notable difference in the statistics of SC and ST population is that while the maximum SC population across districts is 50%, there are many districts with ST population as the majority, particularly the North-eastern states, which is as high as close to 95%.

Vaccine hesitancy had been particularly high globally among the members of the muslim community surrounding their religious beliefs (Dube et al., 2014; Shrivastawa et al., 2019; Khan et al., 2020; Harapan et al., 2021). The vaccination rates across percentage of muslim population in districts do not show any correlation. Several districts in Jammu and Kashmir that had high muslim majority did exceptionally well with covid-19 vaccine coverage (The Print; Hindustan Times). Excluding districts of Jammu and Kashmir, a strong negative correlation of vaccination rates with percent of muslim population in a district can be seen during the third phase of the vaccination drive. Besides the fact that the second covid wave would have caused vaccine hesitancy, these signs of vaccine hesitancy among the muslims could also be due to the month-long Ramadan (fasting month for muslims) between April 13 to May 12, 2021. In spite of clarifications by muslim scholars that vaccine would not invalid the fast, there have been evidence of vaccine hesitancy among muslims (Ali et al., 2021).

The distribution of ST and muslim population across all districts is more variable than the SC population distributions. Unlike the SC population, which is not greater than 50% in all districts, percentage of ST and muslim population is greater than 90% in several districts for example the north-eastern states and Jammu and Kashmir respectively. While the SCs, STs and muslims are constitutionally recognized social and religious minorities respectively, in the north-eastern states STs form the majority and muslims form the majority in several districts of Jammu and Kashmir. Both these regions have relatively higher rates of covid vaccination compared to other districts where STs and muslims are not in a significant majority. Social network and peer effects can highly influence vaccination decisions. With high concentration of a minority in a particular region, it is highly likely that the vaccination drive or any other public programme will involve people from the minority group or the local community heavily which is less likely in regions where the minority sections are marginalised or do not actively participate in public programmes. This could lead to higher confidence and success of public programmes in districts with high concentration of minority communities. However, these are mere conjectures and would require further research to be established as evidence.

Child immunization rates are found to be positively correlated with the covid vaccination rates across all phases except the first phase of vaccination for the frontline workers. Although adult immunization is uncommon in India, child vaccination under the UIP has existed for more than three decades and is popular among the masses. Districts that had higher full child immunization rates also had higher covid vaccination rates. The positive relationship between full child immunization and covid vaccination was even stronger during the third phase of the vaccination drive compared to the second and the fourth phase. As the vaccine hesitancy was the highest during the third phase of the vaccination drive, the above correlation could be indicative of overall attitude towards vaccines and not just covid vaccine. Regions with low immunization rates among the children can be areas that are likely to be potential hotspots for vaccine hesitancy. The positive correlation between district level full child immunization rates and covid vaccination rates are significant across both NFHS-4 and NFHS-5 rounds for the years 2014-15 and 2019-20 respectively.

Several research studies have found that one of the prevalent reasons for vaccine hesitancy has been concerns surrounding fertility. We do not find any overall relationship between district level indicators of family planning and covid vaccination rates. However, during the phase three of the vaccination drive, when vaccine hesitancy was the highest there seems to be a positive relationship between any family planning method adoption rate and covid vaccination rate. Since, female sterilization was the primary family planning method used across all districts a similar relationship between district level female sterilization rate and covid vaccination existed. There exists an inverse relationship between unmet need for family planning and covid vaccination rates at the district level only phase three of the vaccination drive. While family planning indicators might not be correlated with vaccination rates, low performance of these indicators could be regions that could turn out to be hotspots for vaccine hesitancy. There is evidence towards correlation between vaccination rates and family planning indicators across both NFHS-4 and NFHS-5 rounds. However, the correlation is weaker for NFHS-5 rounds. This could be because NFHS-5 does not include data for all states compared to NFHS-4 as the survey was affected due to the pandemic, which delayed the release of the data for districts of all the states. There could be systemic bias in the partial NFHS-5 data used. Alternatively, it is also possible that between the NFHS-4 and NFHS-5, this relationship has weakened. However, nothing concrete can be said unless the full data for the NFHS-5 with all the states is available.

A limitation of our study is that we do not observe vaccine hesitancy directly. We use low vaccination rates as a proxy for vaccine hesitancy. As vaccine hesitancy is defined as refusal or delay in getting vaccinated in spite of availability of a vaccine (MacDonald, 2015), we implicitly assume that the low vaccination rate was not a result of short supply of vaccine. While low vaccination rates could be driven by inadequate vaccine supply, our observations on vaccine hesitancy from district level trends on vaccination uptake would still hold unless there is a compelling reason to believe that vaccine shortages and supply disparities were motivated by socio-economic factors. We are not familiar with evidence that can support a pattern in short supply of vaccines across the districts.

## Conclusion

In our current study, we use district level data from the COVID-19 vaccination uptake in India to discuss the correlates of vaccine hesitancy. We find heterogeneity of vaccine uptake across different districts in India which is correlated with the demographics and socio-economic factors. Past district level child immunization and family planning indicators too are correlated with vaccination uptake rates. Using the phase wise rollout of district level COVID-19 vaccination drive, regions that are potentially most susceptible to vaccine hesitancy have been indicated. As COVID-19 vaccination for children are being developed and are in the pipeline, evidence on spots of vaccine hesitancy from our study could be of huge significance for the COVID-19 vaccination drive in India. Evidence on vaccine uptake and vaccine hesitancy from India’s massive vaccination drive can also serve as an important lesson for COVID-19 vaccination in other LMICs and for better preparedness for future public health outcomes.

## Data Availability

The data used in this study is in public domain. Data sources have been cited in the article.

https://doi.org/10.7910/DVN/RXYJR6

https://doi.org/10.7910/DVN/BL6NMM

## Appendix

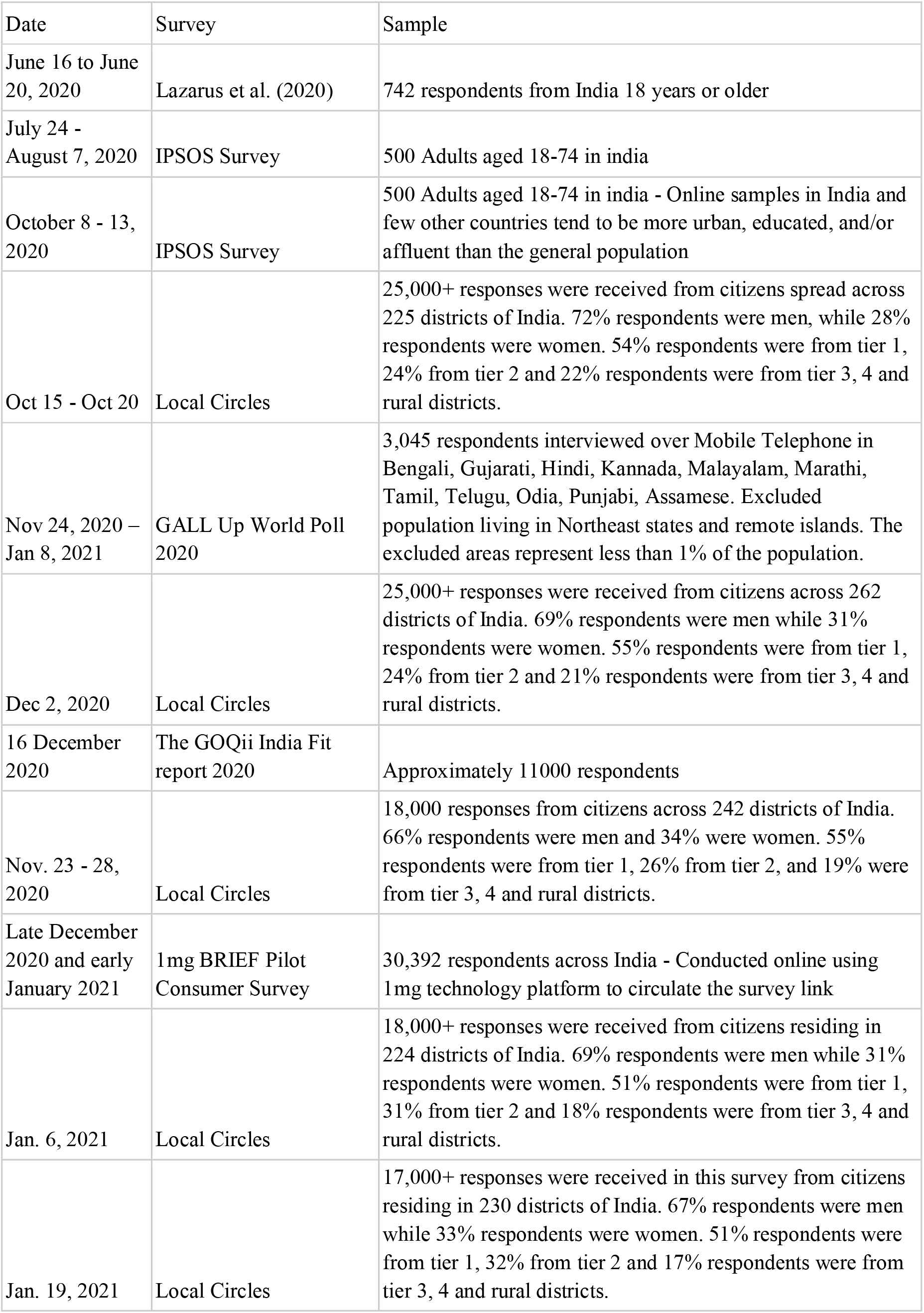
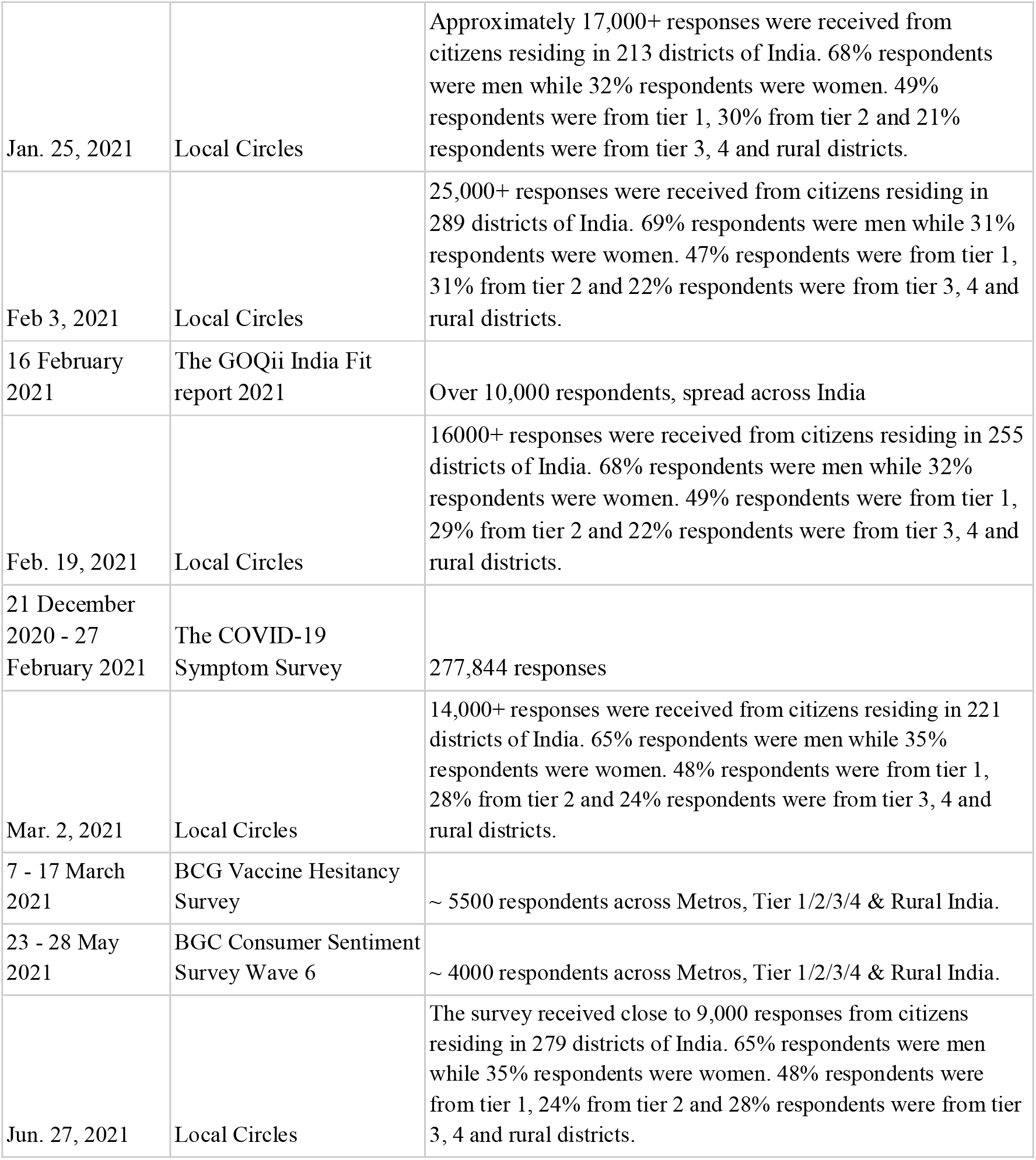

